# Cost and cost-effectiveness of alternative screening and diagnostic pathways for achieving hepatitis C elimination in the country of Georgia

**DOI:** 10.1101/2025.05.22.25327932

**Authors:** Josephine G. Walker, Irina Tskhomelidze Schumacher, Shaun Shadaker, Tamar Gabunia, Rania A. Tohme, Akaki Abutidze, Vladimer Getia, Peter Vickerman

## Abstract

**Background:** We evaluated the cost and cost-effectiveness of alternative screening pathways during 2018–2022 within Georgia’s hepatitis C elimination program, which started in 2015.

**Methods:** We calculated patient-level costs (2022 USD$) of hepatitis C treatment with centralized and decentralized diagnostic testing in hospitals, primary healthcare (PHC), harm reduction providers (HRP), or specialized providers (SP) from reimbursement records including the value of donated direct-acting antivirals (DAAs). We model hepatitis C case-finding, transmission, and progression over a 20-year time horizon to project cost-effectiveness of treatment for each screening pathway in terms of cost per quality adjusted life year (QALY), compared to a willingness-to-pay threshold of $1,337.

**Findings:** Unit costs of treatment decreased from $3942–$4247 across screening pathways in 2018 to $300–$338 in 2022, primarily due to reductions in DAA costs. The cascade of care varied by screening pathway, with highest hepatitis C virus (HCV) antibody prevalence and percent linked to viremia testing among HRP and SP, while total number of patients screened was highest in hospitals. While DAA costs decreased, the cost of case finding increased during 2018–2022, with the biggest increase in hospital settings mainly due to decreasing yield. The program was not cost-effective with full DAA costs, but excluding DAA costs or using lower 2022 costs make all pathways cost-effective and SP, HRP, and PHC potentially cost-saving.

**Interpretation:** Donated drugs allowed Georgia’s HCV elimination program to be cost-effective, while future programs with generic drug costs are likely to be cost-effective. Reductions in yield from hospital screening suggest that later stages of elimination programs should prioritise targeted pathways.

## Introduction

In 2015, a national survey found the prevalence of hepatitis C virus (HCV) infection in Georgia was among the highest in the world. Among adults, 7.7% had antibodies to HCV and 5.4% had chronic hepatitis C, resulting in an estimated 150,000 (95% confidence interval (CI): 128,060 to 173,060) cases.^1^ Alongside the national survey in 2015, Georgia adopted a national hepatitis C elimination program, which aims to achieve the elimination of hepatitis C as a public health threat through implementing a comprehensive screening and treatment program.^2^ The original program goals were to diagnose 90% of infections, treat 95% of those diagnosed, and cure 95% of treated patients by 2020.^3,4^ In addition, the Global Health Sector Strategy to eliminate hepatitis C as a public health threat aims to reduce chronic hepatitis C incidence by 80% and hepatitis C mortality by 65% by 2030.^5^

As of 1 January 2023, over 2.8 million people had been tested at least once for HCV antibodies (anti-HCV) in Georgia, out of a population of 3.7 million. A total of 80,997 people had initiated treatment, with a cure rate of 96.9% among those tested for sustained virological response (SVR) after their first round of treatment. A repeat national sero-survey conducted in 2021, after over 70,000 persons were treated, demonstrated a large decrease in prevalence of chronic hepatitis C to 1.8% among adults (6.8% anti-HCV prevalence); modelling estimates a 58% decrease in incidence of HCV infection during 2015–2022 based on serosurvey prevalence results.^6,7^

A previous study estimated that the early stage of the hepatitis C elimination program, through November 2017, was likely to be cost-effective for the government of Georgia, as direct-acting antivirals (DAAs), which were expensive at that time, were donated for free and continue to be donated to the program.^8^ Since 2020, generic DAAs have been available globally at low cost, indicating that the program, including paying for DAAs, could be cost effective even at a low willingness-to-pay (WTP) threshold. In this study, we aim to calculate the cost per patient treated based on screening observed at different sites, accounting for differences in anti-HCV prevalence and loss to follow up along the care cascade. We explore how the cost of case finding and treatment has changed over time. We then calculate the cost per quality-adjusted life year (QALY) gained for each of the screening strategies during 2018–2022, by projecting the long-term impact of each treatment in a dynamic model of HCV transmission and disease progression in Georgia. Finally, we explore how changes in costs and prevalence yield could affect the cost-effectiveness of the screening strategies.

## Methods

We used government expenditure records and programmatic data on the hepatitis C care cascade from the elimination program within a decision tree model to estimate the cost per patient treated for alternative screening strategies used in Georgia during 2018–2022. We account for differences in prevalence yield in testing and levels of linkage to viremia testing and treatment across screening settings. We then use a dynamic model to estimate the incremental cost-effectiveness ratio for each pathway, in terms of cost per QALY gained, compared to if each strategy was not included in the elimination program. Cost-effectiveness results are compared to a conservative opportunity-cost based WTP threshold of 20% GDP per capita ($1,337 USD) in 2022.^9^

### Screening Strategies

Routine screening for anti-HCV takes place in a variety of settings in Georgia, including both hospital and community-based settings. The five screening strategies evaluated in this study are at 1) centralized hospital, 2) decentralized hospital, 3) primary healthcare (PHC), 4) harm reduction providers (HRPs), and 5) specialized treatment sites (Table 1). Under all pathways, if active infection is diagnosed (detected HCV RNA or positive HCV core antigen), patients are referred for treatment at their preferred location (PHC, HRP, or specialized treatment site).

**Table 1:** Summary of screening sites for hepatitis C, including number of people screened, number of people treated after diagnosis at that screening site, source of anti-HCV rapid tests, and diagnostic tests used, during 2018–2022 in Georgia.

| Screening site | Description | N people screened 2018–2022 | N treatments 2018–2022 | Anti-HCV (Hepatitis C Rapid Test) test source | Diagnostic test used |
| --- | --- | --- | --- | --- | --- |
| Centralized hospital sector | all hospital inpatients were tested for anti-HCV on site, and samples (remaining blood where possible) from anti-HCV positive patients were submitted to the central national reference laboratory (Lugar Center) for viremia testing | 667,124 | 4,233 (0.6%) | Privately purchased | HCV Core Antigen (HCV coreAg), with negative samples and those in the grey zone retested by PCR for HCV RNA to confirm results |
| Decentralized hospital sector | inpatients at participating hospitals were tested for anti-HCV on site, and samples from anti-HCV positive patients were submitted to selected specialized HCV provider labs in Tbilisi, where HCV RNA testing was conducted | 700,212 | 5,632 (0.8%) | Privately purchased | HCV RNA by point of care test (GeneXpert*) or laboratory-based PCR |
| Primary healthcare | rapid tests were used to test for anti-HCV, and positive samples were sent to contractor laboratories for viremia testing using either GeneXpert or standard qualitative or quantitative PCR | 98,182 | 459 (0.5%) | NCDC provided | HCV RNA by point of care test (GeneXpert*) or laboratory-based PCR |
| Harm reduction providers | rapid tests were used to test for anti-HCV, and on-site HCV RNA testing was done using a GeneXpert machine. People tested in this model were generally beneficiaries of the harm reduction service (needle and syringe provision or other services) or their family members | 5,204 | 469 (9.0%) | Privately purchased | HCV RNA by point of care test (GeneXpert*) |
| Specialized provider | anti-HCV testing was performed at specialized HCV treatment facilities, including clinics with a focus on infectious diseases. Some clinics sent anti-HCV positive samples to the Lugar Center for viremia testing, while others performed viremia testing on site using qualitative or quantitative PCR. Since 2018, the majority of tests have been sent to the Lugar Center for viremia coreAg testing | 92,116 | 4,889 (5.3%) | NCDC provided | HCV coreAg |

Abbreviations: HCV, hepatitis C virus, NCDC, National Center for Disease Control and Public Health, anti-HCV, hepatitis C virus antibody; \*the point of care HCV RNA testing system used is GeneXpert

From the launch of the decentralization of hepatitis C care and treatment among PHC and HRP in August 2018 through 2022, a total of 11 PHC and 6 HRP settings throughout the country were providing hepatitis C screening and care services. In the initial phase of decentralization by the government, only HCV treatment-naïve patients with no or mild fibrosis (FIB-4 score <1.45) were treated using simplified diagnostics and treatment monitoring in PHC and HRP sites. Treatment eligibility criteria were later expanded to those with FIB-4 scores between 1.45 and 3.25, while persons with cirrhosis were referred to specialized clinics. Decentralized HRP sites had elastography on site, but PHC referred to specialized sites for elastography if needed.

Overall, more than 97% of treated patients diagnosed through the hospital models or specialized provider were treated by specialized providers. Of those diagnosed in PHC, 35.9% were treated at PHC and the remainder at specialized providers. Of those diagnosed in HRP, 60.8% were treated at HRP, 0.6% at PHC, and the remainder at specialized providers. We used a cut-off date of 31 December 2022 for treatment initiation.

### Costs

Cost data were provided by the Ministry of Internally Displaced Persons from the Occupied Territories, Labour, Health, and Social Affairs of Georgia (hereafter referred to as MoH), National Center for Disease Control and Public Health (NCDC) and service providers. From these data we calculated unit costs for anti-HCV tests, viremia tests (HCV core antigen or HCV RNA), patient-level totals for treatment and monitoring, and cost of care for liver disease. Costs were collected in Georgian Lari (GEL), inflated to 2022 values using the World Bank consumer price index for Georgia, and converted to USD using an average exchange rate for 2022 of 2.915 GEL/USD. Costs are presented from the health system perspective, including both provider (MoH) and patient costs, accounting for patient co-payments where applicable, and the value of donated drugs. Full details of costing methodology for anti-HCV screening tests, viremia tests, treatment and monitoring, DAA costs, and liver disease care are provided in the Supplementary Methods.

### Decision tree model

Each patient was assigned to one of the five screening strategies described above based on their first anti-HCV positive screening result recorded in the elimination program, or most recent negative if no positive results. To account for potential duplicate entries, repeated test results for the same person within 3 days of a previous test were excluded.

For each screening strategy in each year during 2018–2022, the decision tree shown in Figure 1 was used to calculate the total cost per person treated. First, the total diagnosis cost for each year was calculated by multiplying the anti-HCV test cost by number of tests given, added to the viremia test cost multiplied by number of persons tested for viremia within that year. As treatment did not always occur in the same year as screening, and treatment costs changed over time, the treatment cost for each year of screening was calculated as the treatment cost multiplied by the number starting treatment in each year that had been screened in a particular year. Screening numbers by year and when and where they were treated are shown in Supplementary Table 1. The sum of diagnostic and treatment costs was then divided by the number starting treatment to get the annual cost per person treated under each screening strategy.

**Figure 1:**
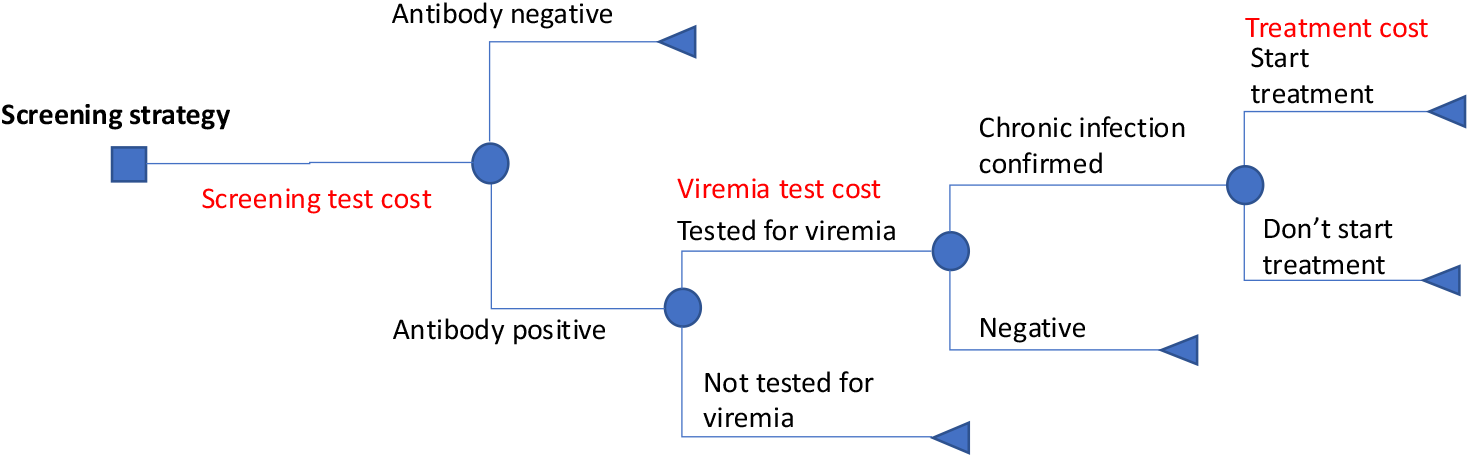
Decision tree model of the hepatitis C diagnostic pathway, used to calculate cost per person treated under each screening strategy

### Dynamic model

We used a previously published and validated dynamic model of HCV transmission in Georgia to project the long term costs and impacts of treatment^7,10^. Briefly, the compartmental model was designed to represent the past and current HCV epidemic in Georgia, accounting for HCV transmission among people who inject drugs (PWID) and the general population. The model is stratified by infection status, sex, age, liver disease status, and injecting drug use. The model was parameterised and calibrated to data from the 2015 and 2021 national sero-surveys and PWID surveys. The model accounts for observed treatment numbers through the end of 2022, with the impact of treatment projected forward for 20 years. For this analysis, the model was adapted and run in Matlab 2023b, with output files analysed in R version 4.4.^11^ We use the set of 76 parameters under the filtered scenario which fit the results of the 2021 serosurvey as described in a previous publication.^7^

### Health-related quality of life

We assigned health-related quality of life utility weights to each modelled liver disease state (mild, moderate, cirrhosis, decompensated cirrhosis, or hepatocellular carcinoma (HCC)).^8,12^ The utility weights and uncertainty ranges were, for mild disease 0.83 (0.81–0.85), for moderate disease 0.81 (0.77–0.85), cirrhosis 0.74 (0.68–0.8), decompensated cirrhosis 0.66 (0.46–0.86), and HCC 0.65 (0.44–0.86).

### Cost-effectiveness analysis

A total of 80,897 patients were treated by the end of 2022, with 38,522 treated during 2018–2022. The number of treated patients identified from each of the screening strategies during 2018–2022 were: 4233 for the centralized hospital sector strategy, 5632 for the decentralized hospital sector strategy, 459 for the PHC strategy, 469 for the HRP strategy, and 4889 for the specialized provider strategy. To estimate the impact in terms of QALYs gained due to each screening strategy, we compared the impact of the full set of treatments through 2022 compared to removing the treatments for each of the models. For simplicity of comparison, HCV treatment is assumed to stop after 2022.

We undertook a probabilistic analysis by sampling 500 values for the model outputs (number in each liver disease state by year) from the selected model parameter sets, with QALY weights sampled from triangular distributions. Total treatment costs per patient (including the cost of case finding from each screening pathway) were sampled from a normal distribution assuming a standard error of 5% of the mean treatment cost for each pathway in each year. Cost of liver disease care was also sampled from a normal distribution using a standard error of 5% of the mean value.

The mean difference in total costs and total QALYs by pathway through 2040 was calculated in comparison to the baseline to obtain the incremental cost-effectiveness ratio (ICER). The ICER is presented in terms of cost per QALY from the health system perspective with a 3% annual discount rate.

We conducted sensitivity analyses on the perspective by (1) excluding donated DAA costs (provider and patient perspective); (2) excluding cost of liver disease and DAAs to show the impact of the program without accounting for averted healthcare costs (elimination program perspective); and (3) we applied 2022 costs for each pathway for all years during 2018–2022, including DAA costs, to represent health system perspective in which a treatment program is started from scratch with lower drug costs. In addition, we conducted a standard sensitivity analysis on the base case health system perspective with 0% discount rate or a shortened time horizon to 2030.

Role of the funding source: This study was funded by Gilead Sciences through an investigator-sponsored research grant to PV and JGW. The funder played no role in the study design, collection, management, analysis, or interpretation of data, in the writing of the report, or in the decision to publish.

## Results

### Unit costs

Unit costs for antibody screening, viremia testing, and treatment within each screening pathway and year are presented in Supplementary Table 2. Treatment costs decreased over time, with a clear shift from 2019 to 2020, due primarily to a reduction in the cost of DAAs (Figure 2). Drug costs then remained relatively stable over 2021–2022, between $227 and $252 per course of treatment in 2022. The non-drug component of treatment costs decreased consistently across all treatment pathways each year. In 2018, HRP settings had the lowest total treatment cost (including DAAs and treatment monitoring) of $3942 while specialized providers had the highest at $4247. In 2022, the HRP pathway still had the lowest treatment cost at $300 while the highest cost was for specialized providers at $338.

**Figure 2:**
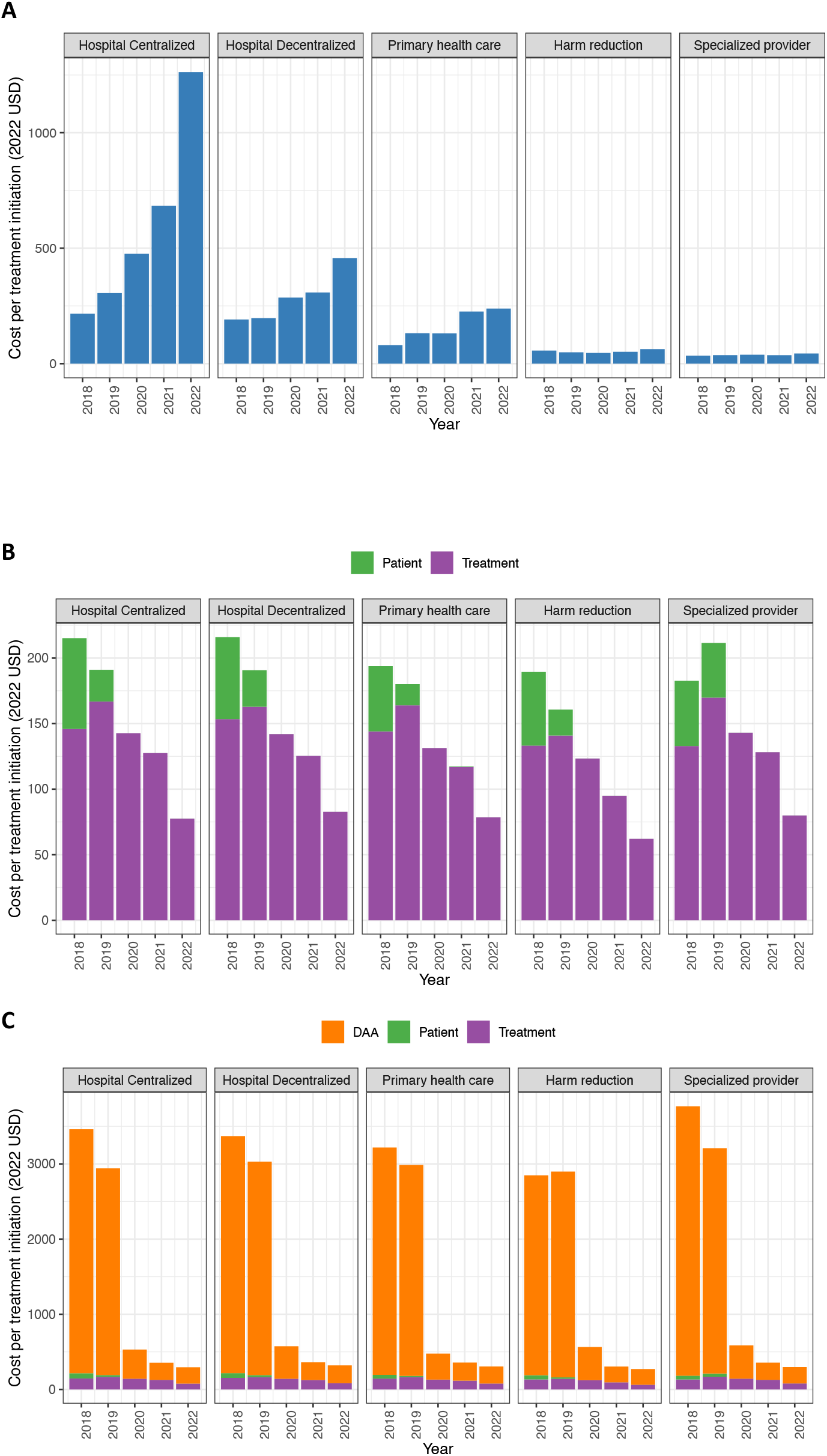
(A) Cost of hepatitis C diagnosis per treatment initiation; (B) Cost of hepatitis C treatment monitoring; and (C) Cost of DAAs with cost of hepatitis C treatment monitoring, for each person treated by screening pathway and year under the hepatitis C elimination program in Georgia, 2018–2022. Green bars are patient out of pocket costs during treatment, purple bars are costs to elimination program during treatment, orange bars DAA costs. DAAs: direct-acting antivirals.

A total of 886 persons in 2015 and 770 persons in 2016 were included in the financial database for liver disease care; 273 people had costs recorded in both years. The cost per person treated per year was $280.58 for moderate liver disease, $420.08 for compensated cirrhosis, and $830.49 for decompensated cirrhosis, which we assumed applied to HCC.

### Screening strategies

The number of patients included in each screening model and the cascade of care for hepatitis C through treatment outcome are shown in Table 2. Hospital-based screening reached the most patients for anti-HCV testing, with both the centralized and decentralized strategies conducting approximately 1 million anti-HCV tests over 2018–2022, compared to fewer than 120,000 tests in PHC and specialized provider sites, and 5,505 tests in HRP. However, hospital-based testing was more likely to be repeated, with 1.57 tests per unique person in centralized and 1.38 tests per person in decentralized strategies, compared to 1.06–1.29 in the other strategies. HRP, followed by specialized providers, had the highest anti-HCV positivity rate among persons tested, with 17.7% and 8.8% respectively, compared to less than 3% in the other settings. These two strategies also had higher rates of viremia testing among those that were anti-HCV positive compared to hospital and primary healthcare. The proportion of anti-HCV positive patients with chronic HCV infection was also lower in hospital settings, which could be due to re-testing of patients who had already been treated. Specialized providers were the most likely to link diagnosed patients to the treatment program (87.9%), followed by HRP (77.7%) and PHC (73.6%), with much lower rates of linkage to care among persons diagnosed in hospital settings (55.0–64.7%). However, once patients were enrolled in the hepatitis C treatment program, rates of starting (93–95%) and completing (90–94%) treatment and of achieving SVR (97–99%) were similar across all strategies.

**Table 2:** Cascades of care for each hepatitis C screening site in Georgia. The number of persons at each step in the care cascade with percent who completed each step compared to the number in the previous row.

|  | Hospital Centralized<br>(2018–2022) |  | Hospital<br>Decentralized |  | Primary Healthcare |  | Harm reduction<br>providers |  | Specialized Provider<br>(2018–2022) |  |
| --- | --- | --- | --- | --- | --- | --- | --- | --- | --- | --- |
| **Tests Conducted\*** | 1,047,456 |  | 969,697 |  | 119,186 |  | 5,505 |  | 118,460 |  |
| **Unique Persons Screened** | 667,124 | 1.57 TPP | 700,212 | 1.38 TPP | 98,182 | 1.21 TPP | 5,204 | 1.06 TPP | 92,116 | 1.29 TPP |
| **Anti-HCV Positive\*** | 15,469 | 2.3% | 18,146 | 2.6% | 1,173 | 1.2% | 923 | 17.7% | 8,123 | 8.8% |
| **Tested for Viremia\*** | 11,734 | 75.9% | 14,434 | 79.5% | 811 | 69.1% | 812 | 88.0% | 7,494 | 92.3% |
| **Chronic HCV\* Infection** | 8,192 | 69.8% | 9,338 | 64.7% | 648 | 79.9% | 649 | 79.9% | 5,844 | 78.0% |
| **Enrolled in Program** | 4,502 | 55.0% | 6,040 | 64.7% | 477 | 73.6% | 504 | 77.7% | 5,134 | 87.9% |
| **Started Treatment\*** | 4,233 | 94.0% | 5,632 | 93.2% | 459 | 96.2% | 469 | 93.1% | 4,889 | 95.2% |
| **Completed Treatment** | 3,895 | 92.0% | 5,184 | 92.0% | 430 | 93.7% | 422 | 90.0% | 4,558 | 93.2% |
| **Eligible for SVR** | 3,750 | 96.3% | 4,891 | 94.3% | 419 | 97.4% | 403 | 95.5% | 4,413 | 96.8% |
| **Tested for SVR** | 2,823 | 75.3% | 3,279 | 67.0% | 323 | 77.1% | 300 | 74.4% | 3,177 | 72.0% |
| **SVR Achieved** | 2,785 | 98.7% | 3,238 | 98.7% | 313 | 96.9% | 292 | 97.3% | 3,115 | 98.0% |

Abbreviations: TPP, tests per person; HCV, hepatitis C virus, anti-HCV, HCV antibody; SVR, sustained virologic response. **\*indicates steps represented in the decision tree model (Figure 1).**

### Cost per diagnosis and treatment

Case finding costs are driven by changes in yield of positive test results, which decrease over time in hospital settings. The diagnostic costs (including anti-HCV and viremia testing) per treatment initiation increased from $216 in 2018 to $1262 in 2022 in the centralized hospital strategy, and from $191 to $456 over the same period in the decentralized hospital strategy. PHC costs also increased from $80 to $239, while HRP and specialized provider diagnosis costs were relatively stable and between $34 to $63 per patient treated (Figure 2A).

The total costs per treatment initiation based on treatment monitoring alone within each strategy and excluding diagnostic costs also decreased over time in all settings, with costs fairly similar across pathways (Figure 2B). When DAA costs are accounted for, there is a large decrease in treatment cost from 2019 to 2020 due to the switch to generic regimens followed by a gradual decrease reaching $300–$338 per treatment initiation in 2022 (Figure 2C). The cost of treatment accounts for actual year of treatment initiation (for example, 50% of those screened in 2018 were treated in 2018) which results in the cost per treatment initiation by pathway being lower than the unit costs by year in some cases (Supplementary Tables 1 and 2).

Overall, the cost of DAA drugs has changed from being the primary driver of treatment costs to being similar or less than the cost of case finding diagnostics (Figure 2). Across all years, HRP had the lowest cost per treatment initiation, due to high yield and slightly lower treatment costs (Table 3). Based on more recent 2022 costs applied to the cascade of care and including DAA costs, the cost per treatment initiation is lowest in HRP ($350) followed by specialized providers ($375), with PHC $423, hospital decentralized and hospital centralized the highest at $589 and $681, respectively, per patient treated.

**Table 3:** Total cost per hepatitis C treatment initiation in Georgia, by screening pathway and year of screening, in 2022 USD, and total QALY losses averted by treatments under each screening pathway.

| Year | Hospital Centralized | Hospital Decentralized | Primary Healthcare | Harm reduction providers | Specialized Provider |
| --- | --- | --- | --- | --- | --- |
| Total cost including DAAs (health system perspective) |  |  |  |  |  |
| 2018 | 3617 | 3480 | 3228 | 3179 | 3844 |
| 2019 | 3126 | 3057 | 3008 | 2821 | 3080 |
| 2020 | 1602 | 1399 | 1116 | 967 | 1294 |
| 2021 | 1052 | 683 | 588 | 388 | 412 |
| 2022 | 1591 | 786 | 544 | 363 | 381 |
| Average over 2018-2022 | 1861 | 1319 | 1491 | 1012 | 1512 |
| Average over 2018-2022 using 2022 costs | 681 | 589 | 423 | 350 | 375 |
| Excluding DAA costs (provider and patient perspective) |  |  |  |  |  |
| 2018 | 425 | 393 | 270 | 231 | 261 |
| 2019 | 490 | 388 | 311 | 211 | 236 |
| 2020 | 623 | 435 | 271 | 171 | 195 |
| 2021 | 806 | 434 | 345 | 152 | 166 |
| 2022 | 1349 | 545 | 315 | 135 | 130 |
| Overall | 280 | 221 | 157 | 97 | 118 |
| Overall using 2022 costs | 438 | 348 | 193 | 122 | 123 |
| QALY losses averted through 2040 by treatments under base case assumptions |  |  |  |  |  |
|  | 1975 | 2310 | 2257 | 163 | 187 |

Abbreviations: QALY, quality adjusted life years; DAA, direct-acting antiviral

### Cost-effectiveness

Under the base case scenario over a time horizon to the end of 2040, and including liver disease costs and the cost of DAA drugs, none of the pathways are cost effective at the WTP threshold of $1,337/QALY but are likely to be cost-effective at a higher threshold of 1 GDP per capita ($6,685) (Supplementary Table 3, Supplementary Figure 1). However, when DAA costs are removed (provider plus patient perspective), all screening pathways are cost effective at the lower threshold, with specialized provider, HRP, and PHC strategies being cost saving due to higher screening yields (Table 4, Supplementary Figure 1). Excluding liver disease costs as well as DAAs results in the mean ICER for all scenarios remaining cost-effective, with 76.6%, 86.0%, and 99.8% of model runs cost-effective for centralized, decentralized, and PHC strategies, respectively, and all model runs cost-effective for specialized providers and HR strategies. Using 2022 costs for the whole care pathway, and including DAA costs, all ICERs are below the WTP threshold with the mean ICER for specialized providers, HRP, and PHC all cost-saving. A shorter time horizon to 2030 means the strategies are no longer cost-effective due to the up-front cost of the treatments and longer time horizon for QALY gains. Without discounting, all ICERs are lower compared to the base case, falling above 20% GDP per capita but below 1 GDP per capita.

**Table 4:**
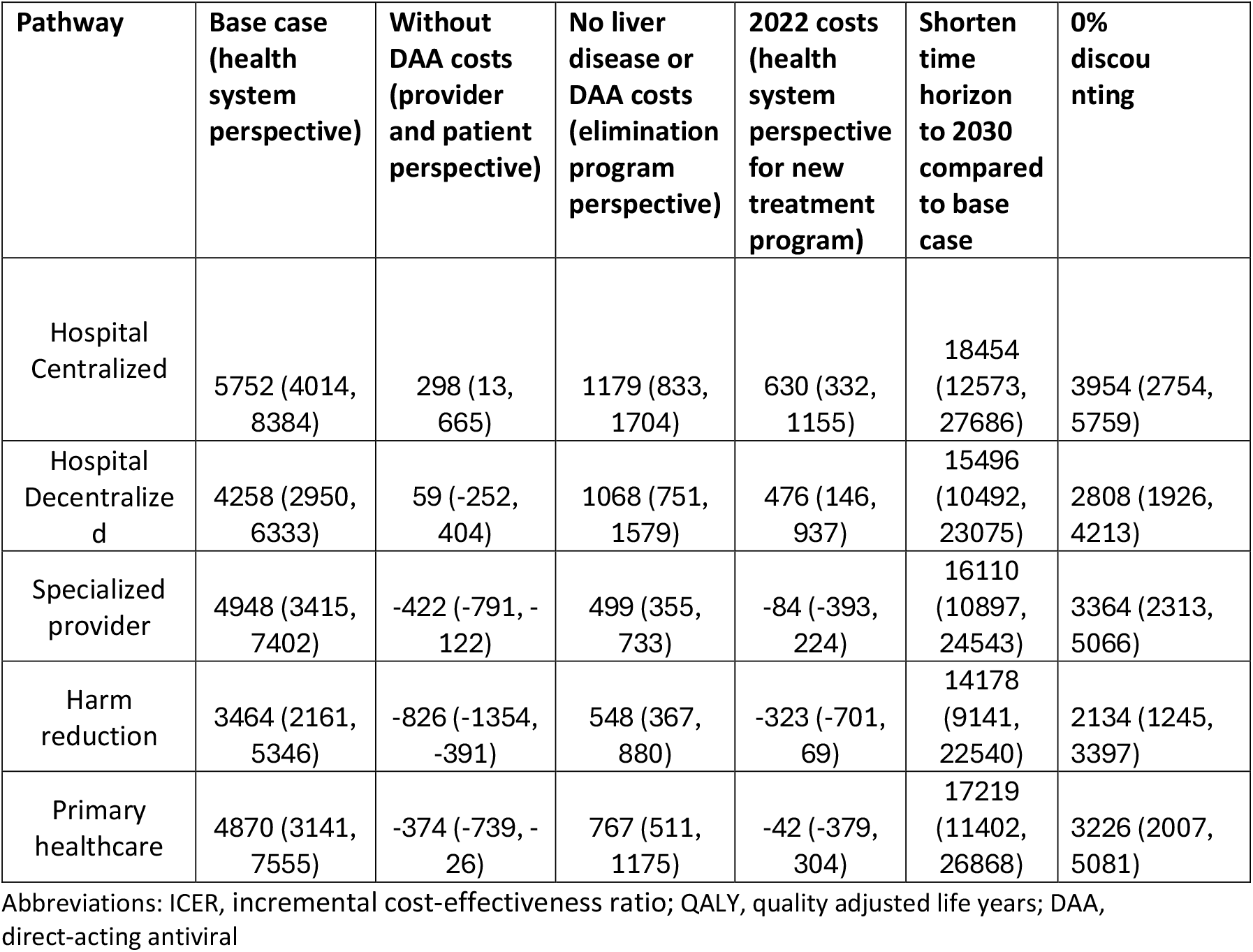
Mean incremental cost-effectiveness ratio (ICER) in terms of cost per quality adjusted life year (QALY) in 2022 US dollars with a time horizon to 2040, and 95% credible interval for each hepatitis C screening pathway compared to baseline, for base case model assumptions and scenario analyses.

## Discussion

Our results highlight significant reductions in the cost of HCV treatment during 2018–2022 across various diagnostic strategies in Georgia, driven by changes in screening yield, the introduction of generic drugs, and simplification of treatment protocols. We find that from the provider and patient perspective, with DAA drugs donated to the program throughout, the elimination program across all evaluated pathways is likely to have been cost-effective, and may have been cost-saving for screening pathways outside of hospitals due to averted liver disease care costs. The same program implemented based on 2022 costs including DAA costs would similarly be cost-effective or cost-saving, indicating that DAA costs are less of a barrier to hepatitis C elimination than they were when elimination targets were introduced in 2016.^13,14^

We identified clear differences in the cost-effectiveness of different screening pathways, with screening in HRPs and PHCs, as well as specialized providers leading to higher yield, with better linkage to care and treatment uptake. In Georgia, HCV screening is mandatory for all hospital inpatients, and early in the elimination program this allowed identification of persons with HCV infection without previously identified risks of HCV infection. However, over time the screening yield and efficiency of the diagnostic pathway decreased in hospital settings, while it was maintained by the providers who use a more targeted approach for screening. This change over time emphasizes the need for different approaches as fewer persons remain to be diagnosed and linked to care.

### Strengths and limitations

A key strength of our study is the use of comprehensive, real-world data on costs and care cascades. We also build on robust modelling techniques that provide a detailed analysis of costs and outcomes. However, real-world data introduced some challenges and limitations around developing the care cascade and we had limited data to support some assumptions. Patient pathways were complex, with many patients being screened in one setting but treated in another, or with a long delay between diagnosis and treatment. We were able to capture this within our cost estimates by focusing our analysis on cohorts defined by their screening site (where they had their first positive test) and accounting for costs of treatment for persons wherever and whenever they were treated. Although this loses nuance in the cascade of care that may depend on where patients enroll in the program or decide to be treated, it accounts for the reality of when and where people were treated. In addition, the total number of people reached through harm reduction is an underestimate, as some patients are tested anonymously in these settings and therefore were not linkable across the care cascade.^4^

Some costs also had to be simplified, and we assumed a normal distribution for costs. We did not estimate the staff time or overhead associated with screening tests as no data were available on this. For HCV RNA costs, test type was not specified in the financial database, so we had to assume all PCR tests, whether point of care (GeneXpert) or Qualitative laboratory-based tests, had the same cost. We were able to triangulate HCV core Antigen (coreAg) test numbers with laboratory records to account for required PCR re-testing of “grey zone” results in which the coreAg test is inconclusive. For the cost-effectiveness analysis, the impact of treatment was evaluated compared to the full treatment numbers achieved in the elimination program; that is, we removed the number of treatments achieved from each strategy to calculate the incremental benefits of those treatments. Evaluating the impact of the treatment numbers achieved by a particular strategy as the only treatments modelled (in the absence of other treatments given) would likely give a different result with higher impact per treatment.

### Comparison to other literature

Cost-effectiveness of HCV screening approaches for elimination have been evaluated in other settings around the world, including high income (Ireland^15^, Canada/USA^16^, Spain^17^, Italy^18^, Germany^19^, South Korea^20^), upper-middle income (China^21^, Iraq^22^), and lower-middle income (Pakistan^23^, Cameroon/Cote d’Ivoire/Senegal^24^) countries. These studies generally present hypothetical scenarios either focused on diagnostic approaches or on which populations to target for screening, and consider the impact of starting large scale screening or treatment programs. The study in China is the only one to consider how optimal targeted screening approaches will change as hepatitis C prevalence decreases.^21^ As Georgia implemented a national hepatitis C elimination program earlier than many other countries, our assessment of the real-life differences between screening strategies should prove useful to other countries as they develop strategies.

Previous studies have evaluated components of the cost-effectiveness of screening and treatment for hepatitis C. Two studies compared the cost-effectiveness of alternative diagnostic strategies where HCV coreAg or HCV RNA is used for the viremia test. Sadeghimehr et al found that antibody followed by HCV coreAg testing was the cheapest option and likely the most cost-effective option, though using antibody followed by HCV RNA instead led to more QALYs gained.^25^ Their unit costs were gathered from literature and experts within Georgia’s hepatitis C elimination program and their model considered disease progression within persons with HCV. Adee et al also modelled disease progression with a Markov model, but diagnostic costs were based on a trial and healthcare costs were adapted from USA data; the most cost-effective approach used on-site RNA tests and Fibroscan for liver disease staging.^26^ Georgia was one country included in a model of the cost-effectiveness (in terms of cost per person cured) of introducing HCV self-testing in addition to standard of care.^27^ Therefore this study, which retrospectively evaluates the real-life implementation of different screening approaches within Georgia’s elimination program, and projects long term outcomes within a dynamic model, provides a more complete picture to inform Georgia’s policies as they reach the later stages of elimination. In comparison to our previous paper evaluating the first stages of HCV treatment within the program,^8^ this study demonstrates how the effectiveness and cost-effectiveness of case-finding approaches will change as the number of people already treated increases, but similarly emphasises that the elimination program would not be cost effective while accounting for the full cost of DAAs prior to the drastic decrease in DAA costs seen in recent years.

### Implications

Our study supports the strategic use of a variety of screening and treatment strategies as cost-effective approaches for hepatitis C elimination in Georgia. Although multiple approaches will be needed to reach different groups, effectiveness will change over time. Despite drastic reductions in the cost of treatment, the reduction in yield from mass screening diminishes, thus reducing its cost effectiveness compared to more targeted approaches. This observation is crucial for policy-makers, as it provides evidence that a pivot towards more targeted screening strategies may be necessary in mid to late stages of hepatitis C elimination programs. Furthermore, comparative cost-effectiveness of screening and treatment approaches in Georgia may be particularly relevant for other countries in the Eastern Europe and Central Asia region, where financial constraints and high hepatitis C prevalence often coexist, emphasizing the importance of using efficient care pathways alongside access to affordable DAAs.

## Data Availability

All data produced in the present study are available upon reasonable request to the authors

## Author contributions

JGW, conceptualization, methodology, software, writing – original draft, visualisation, funding acquisition

ITS, conceptualization, writing – original draft, data curation

SS, formal analysis, data curation, writing – review and editing,

TG, project administration, resources

RAT, supervision, project administration, writing – review and editing

AA, conceptualization, investigation

VG, data curation, project administration

PV, supervision, methodology, writing – review and editing, funding acquisition

