## Supplementary Tables and Figures for "Cost and cost-effectiveness of alternative screening and diagnostic pathways for achieving hepatitis C elimination in the country of Georgia"

**Supplementary Table 1: Hepatitis C Care cascades by screening model and year, including treatment sites and years of treatment in Georgia. The number of persons at each step in the care cascade with percent who completed each step compared to the number in the previous row.**

| Screening year 2018 | | | | | | | | | | |
|---|---|---|---|---|---|---|---|---|---|---|
| | Hospital Centralized | | Hospital Decentralized | | PHC | | HR | | Specialized provider | |
| **Care Cascade** | | | | | | | | | | |
| Tests Conducted | 320,842 | | 48,579 | | 13,644 | | 336 | | 48,987 | |
| Unique Persons Screened | 281,503 | | 47,274 | | 13,229 | | 336 | | 41,862 | |
| Anti-HCV Positive | 7,298 | 2.6% | 1,209 | 2.6% | 283 | 2.1% | 48 | 14.3% | 3,236 | 7.7% |
| Tested for Viremia | 6,026 | 82.6% | 1,055 | 87.3% | 214 | 75.6% | 45 | 93.8% | 2,972 | 91.8% |
| Current HCV Infection | 4,283 | 71.1% | 734 | 69.6% | 174 | 81.3% | 36 | 80.0% | 2,360 | 79.4% |
| Enrolled in Program | 2,360 | 55.1% | 452 | 61.6% | 124 | 71.3% | 29 | 80.6% | 2,018 | 85.5% |
| Started Treatment | 2,255 | 95.6% | 429 | 94.9% | 120 | 96.8% | 25 | 86.2% | 1,962 | 97.2% |
| Completed Treatment | 2,089 | 92.6% | 401 | 93.5% | 115 | 95.8% | 22 | 88.0% | 1,840 | 93.8% |
| Eligible for SVR | 2,050 | 98.1% | 395 | 98.5% | 115 | 100.0% | 22 | 100.0% | 1,828 | 99.3% |
| Tested for SVR | 1,545 | 75.4% | 296 | 74.9% | 82 | 71.3% | 17 | 77.3% | 1,329 | 72.7% |
| SVR Achieved | 1,517 | 98.2% | 289 | 97.6% | 79 | 96.3% | 17 | 100.0% | 1,301 | 97.9% |
| **Treating Site** | | | | | | | | | | |
| Specialized Provider | 2196 | 97.4% | 414 | 96.5% | 85 | 70.8% | 17 | 68.0% | 1946 | 99.2% |
| PHC | 38 | 1.7% | 4 | 0.9% | 35 | 29.2% | | | 4 | 0.2% |
| HRP | 21 | 0.9% | 11 | 2.6% | | | 8 | 32.0% | 12 | 0.6% |
| **Year Treated** | | | | | | | | | | |
| 2018 | 957 | 42.4% | 35 | 8.2% | 37 | 30.8% | 5 | 20.0% | 1362 | 69.4% |
| 2019 | 816 | 36.2% | 314 | 73.2% | 54 | 45.0% | 15 | 60.0% | 403 | 20.5% |
| 2020 | 274 | 12.2% | 41 | 9.6% | 18 | 15.0% | 4 | 16.0% | 109 | 5.6% |
| 2021 | 86 | 3.8% | 12 | 2.8% | 7 | 5.8% | | | 46 | 2.3% |

| | | | | | | | | | | |
|---|---|---|---|---|---|---|---|---|---|---|
| 2022 | 122 | 5.4% | 27 | 6.3% | 4 | 3.3% | 1 | 4.0% | 42 | 2.1% |
| **Screening year 2019** | | | | | | | | | | |
| **Care Cascade** | | | | | | | | | | |
| Tests Conducted | 218,268 | | 302,320 | | 36,762 | | 2,427 | | 26,479 | |
| Unique Persons Screened | 193,293 | | 261,127 | | 35,284 | | 2,381 | | 21,430 | |
| Anti-HCV Positive | 3,001 | 1.6% | 6,751 | 2.6% | 473 | 1.3% | 323 | 13.6% | 2,139 | 10.0% |
| Tested for Viremia | 2,497 | 83.2% | 5,714 | 84.6% | 337 | 71.2% | 283 | 87.6% | 2,033 | 95.0% |
| Current HCV Infection | 1,789 | 71.6% | 3,818 | 66.8% | 266 | 78.9% | 231 | 81.6% | 1,564 | 76.9% |
| Enrolled in Program | 1,087 | 60.8% | 2,620 | 68.6% | 192 | 72.2% | 192 | 83.1% | 1,411 | 90.2% |
| Started Treatment | 1,007 | 92.6% | 2,477 | 94.5% | 185 | 96.4% | 186 | 96.9% | 1,342 | 95.1% |
| Completed Treatment | 955 | 94.8% | 2,346 | 94.7% | 178 | 96.2% | 180 | 96.8% | 1,298 | 96.7% |
| Eligible for SVR | 940 | 98.4% | 2,309 | 98.4% | 177 | 99.4% | 179 | 99.4% | 1,291 | 99.5% |
| Tested for SVR | 709 | 75.4% | 1,554 | 67.3% | 146 | 82.5% | 135 | 75.4% | 947 | 73.4% |
| SVR Achieved | 704 | 99.3% | 1,538 | 99.0% | 140 | 95.9% | 130 | 96.3% | 930 | 98.2% |
| **Treating Site** | | | | | | | | | | |
| Specialized Provider | 980 | 97.3% | 2432 | 98.2% | 111 | 60.0% | 86 | 46.2% | 1334 | 99.4% |
| PHC | 26 | 2.6% | 22 | 0.9% | 74 | 40.0% | 3 | 1.6% | 2 | 0.1% |
| HRP | 1 | 0.1% | 23 | 0.9% | | | 97 | 52.2% | 6 | 0.4% |
| **Year Treated** | | | | | | | | | | |
| 2019 | 608 | 60.4% | 1607 | 64.9% | 127 | 68.6% | 129 | 69.4% | 968 | 72.1% |
| 2020 | 298 | 29.6% | 642 | 25.9% | 40 | 21.6% | 46 | 24.7% | 320 | 23.8% |
| 2021 | 42 | 4.2% | 110 | 4.4% | 14 | 7.6% | 6 | 3.2% | 29 | 2.2% |
| 2022 | 59 | 5.9% | 118 | 4.8% | 4 | 2.2% | 5 | 2.7% | 25 | 1.9% |
| **Screening year 2020** | | | | | | | | | | |
| **Care Cascade** | | | | | | | | | | |
| Tests Conducted | 166,780 | | 220,722 | | 26,256 | | 929 | | 17,198 | |
| Unique Persons Screened | 150,457 | | 193,837 | | 24,552 | | 920 | | 14,243 | |
| Anti-HCV Positive | 1,843 | 1.2% | 3,934 | 2.0% | 204 | 0.8% | 182 | 19.8% | 1,105 | 7.8% |

| | | | | | | | | | | |
|---|---|---|---|---|---|---|---|---|---|---|
| Tested for Viremia | 1,345 | 73.0% | 3,128 | 79.5% | 148 | 72.5% | 161 | 88.5% | 1,024 | 92.7% |
| Current HCV Infection | 930 | 69.1% | 1,895 | 60.6% | 113 | 76.4% | 119 | 73.9% | 770 | 75.2% |
| Enrolled in Program | 503 | 54.1% | 1,237 | 65.3% | 92 | 81.4% | 102 | 85.7% | 696 | 90.4% |
| Started Treatment | 476 | 94.6% | 1,166 | 94.3% | 87 | 94.6% | 100 | 98.0% | 655 | 94.1% |
| Completed Treatment | 443 | 93.1% | 1,118 | 95.9% | 83 | 95.4% | 98 | 98.0% | 636 | 97.1% |
| Eligible for SVR | 422 | 95.3% | 1,074 | 96.1% | 81 | 97.6% | 95 | 96.9% | 631 | 99.2% |
| Tested for SVR | 323 | 76.5% | 726 | 67.6% | 61 | 75.3% | 73 | 76.8% | 451 | 71.5% |
| SVR Achieved | 318 | 98.5% | 717 | 98.8% | 60 | 98.4% | 71 | 97.3% | 441 | 97.8% |
| **Treating Site** | | | | | | | | | | |
| Specialized Provider | 457 | 96.0% | 1148 | 98.5% | 57 | 65.5% | 33 | 33.0% | 648 | 99.1% |
| PHC | 15 | 3.2% | 11 | 0.9% | 30 | 34.5% | | | 4 | 0.6% |
| HRP | 4 | 0.8% | 7 | 0.6% | | | 67 | 67.0% | 2 | 0.3% |
| **Year Treated** | | | | | | | | | | |
| 2020 | 302 | 63.4% | 840 | 72.0% | 63 | 72.4% | 77 | 77.0% | 549 | 83.8% |
| 2021 | 114 | 23.9% | 209 | 17.9% | 19 | 21.8% | 16 | 16.0% | 84 | 12.8% |
| 2022 | 60 | 12.6% | 117 | 10.0% | 5 | 5.7% | 7 | 7.0% | 22 | 3.4% |
| **Screening year 2021** | | | | | | | | | | |
| **Care Cascade** | | | | | | | | | | |
| Tests Conducted | 156,044 | | 196,135 | | 22,401 | | 864 | | 11,812 | |
| Unique Persons Screened | 139,546 | | 176,117 | | 21,458 | | 857 | | 11,344 | |
| Anti-HCV Positive | 1,685 | 1.2% | 3,299 | 1.9% | 106 | 0.5% | 180 | 21.0% | 847 | 7.5% |
| Tested for Viremia | 1,123 | 66.6% | 2,589 | 78.5% | 56 | 52.8% | 165 | 91.7% | 774 | 91.4% |
| Current HCV Infection | 683 | 60.8% | 1,612 | 62.3% | 47 | 83.9% | 142 | 86.1% | 613 | 79.2% |
| Enrolled in Program | 338 | 49.5% | 1,027 | 63.7% | 37 | 78.7% | 100 | 70.4% | 558 | 91.0% |
| Started Treatment | 307 | 90.8% | 941 | 91.6% | 36 | 97.3% | 88 | 88.0% | 523 | 93.7% |
| Completed Treatment | 283 | 92.2% | 888 | 94.4% | 34 | 94.4% | 84 | 95.5% | 492 | 94.1% |
| Eligible for SVR | 267 | 94.3% | 844 | 95.0% | 34 | 100.0% | 83 | 98.8% | 484 | 98.4% |
| Tested for SVR | 204 | 76.4% | 553 | 65.5% | 27 | 79.4% | 55 | 66.3% | 342 | 70.7% |

| | | | | | | | | | | |
|---|---|---|---|---|---|---|---|---|---|---|
| SVR Achieved | 204 | 100.0% | 546 | 98.7% | 27 | 100.0% | 54 | 98.2% | 336 | 98.2% |
| **Treating Site** | | | | | | | | | | |
| Specialized Provider | 301 | 98.0% | 931 | 98.9% | 20 | 55.6% | 22 | 25.0% | 522 | 99.8% |
| PHC | 5 | 1.6% | 7 | 0.7% | 16 | 44.4% | | | 1 | 0.2% |
| HRP | 1 | 0.3% | 3 | 0.3% | | | 66 | 75.0% | | |
| **Year Treated** | | | | | | | | | | |
| 2021 | 195 | 63.5% | 672 | 71.4% | 29 | 80.6% | 65 | 73.9% | 430 | 82.2% |
| 2022 | 112 | 36.5% | 269 | 28.6% | 7 | 19.4% | 23 | 26.1% | 93 | 17.8% |
| **Screening year 2022** | | | | | | | | | | |
| **Care Cascade** | | | | | | | | | | |
| Tests Conducted | 185,522 | | 201,941 | | 20,123 | | 949 | | 13,984 | |
| Unique Persons Screened | 165,626 | | 178,598 | | 18,622 | | 937 | | 13,396 | |
| Anti-HCV Positive | 1,642 | 1.0% | 2,953 | 1.7% | 107 | 0.6% | 190 | 20.3% | 796 | 5.9% |
| Tested for Viremia | 743 | 45.2% | 1,948 | 66.0% | 56 | 52.3% | 158 | 83.2% | 691 | 86.8% |
| Current HCV Infection | 507 | 68.2% | 1,279 | 65.7% | 48 | 85.7% | 121 | 76.6% | 537 | 77.7% |
| Enrolled in Program | 214 | 42.2% | 704 | 55.0% | 32 | 66.7% | 81 | 66.9% | 451 | 84.0% |
| Started Treatment | 188 | 87.9% | 619 | 87.9% | 31 | 96.9% | 70 | 86.4% | 407 | 90.2% |
| Completed Treatment | 125 | 66.5% | 188 | 30.4% | 20 | 64.5% | 38 | 54.3% | 292 | 71.7% |
| Eligible for SVR | 71 | 56.8% | 269 | 143.1% | 12 | 60.0% | 24 | 63.2% | 179 | 61.3% |
| Tested for SVR | 42 | 59.2% | 150 | 55.8% | 7 | 58.3% | 20 | 83.3% | 108 | 60.3% |
| SVR Achieved | 42 | 100.0% | 148 | 98.7% | 7 | 100.0% | 20 | 100.0% | 107 | 99.1% |
| **Treating Site** | | | | | | | | | | |
| Specialized Provider | 186 | 98.9% | 617 | 99.7% | 21 | 67.7% | 23 | 32.9% | 405 | 99.5% |
| PHC | 1 | 0.5% | 2 | 0.3% | 10 | 32.3% | | | | |
| HRP | 1 | 0.5% | | | | | 47 | 67.1% | 2 | 0.5% |
| **Year Treated** | | | | | | | | | | |
| 2022 | 188 | 100.0% | 619 | 100.0% | 31 | 100.0% | 70 | 100.0% | 407 | 100.0% |

Abbreviations: PHC, primary health care; HRP, harm reduction provider; SVR , sustained virological response; HCV, hepatitis C virus

**Supplementary Table 2: Mean unit costs for hepatitis C diagnosis and treatment in Georgia per patient by screening pathway and year, in 2022 USD.**

| Year | Screening pathway | Anti-HCV test cost | Viremia test cost | Treatment monitoring cost | DAA cost |
|---|---|---|---|---|---|
| 2018 | Centralized hospital sector | 1.20 | 16.95 | 244.32 | 3963.73 |
| 2018 | Decentralized hospital sector | 1.20 | 22.45 | 246.19 | 3981.69 |
| 2018 | Primary health care | 0.35 | 22.45 | 220.75 | 3811.08 |
| 2018 | Harm reduction | 1.20 | 22.45 | 216.63 | 3725.40 |
| 2018 | Specialized provider | 0.35 | 16.95 | 241.70 | 4005.64 |
| 2019 | Centralized hospital sector | 1.20 | 18.26 | 208.22 | 3638.59 |
| 2019 | Decentralized hospital sector | 1.20 | 21.96 | 214.44 | 3581.87 |
| 2019 | Primary health care | 0.46 | 21.96 | 197.61 | 3556.62 |
| 2019 | Harm reduction | 1.20 | 21.96 | 177.85 | 3400.73 |
| 2019 | Specialized provider | 0.46 | 18.26 | 215.91 | 3511.98 |
| 2020 | Centralized hospital sector | 1.20 | 19.41 | 161.65 | 1401.22 |
| 2020 | Decentralized hospital sector | 1.20 | 21.89 | 158.73 | 1242.85 |
| 2020 | Primary health care | 0.31 | 21.89 | 148.51 | 1075.00 |
| 2020 | Harm reduction | 1.20 | 21.89 | 132.09 | 962.98 |
| 2020 | Specialized provider | 0.31 | 19.41 | 161.65 | 1263.65 |
| 2021 | Centralized hospital sector | 1.20 | 19.96 | 143.95 | 246.58 |
| 2021 | Decentralized hospital sector | 1.20 | 21.02 | 140.80 | 208.75 |
| 2021 | Primary health care | 0.31 | 21.02 | 129.59 | 245.88 |
| 2021 | Harm reduction | 1.20 | 21.02 | 110.68 | 238.87 |
| 2021 | Specialized provider | 0.31 | 19.96 | 139.40 | 244.13 |
| 2022 | Centralized hospital sector | 1.20 | 19.55 | 86.45 | 242.88 |
| 2022 | Decentralized hospital sector | 1.20 | 20.58 | 88.85 | 240.82 |
| 2022 | Primary health care | 0.31 | 20.58 | 76.16 | 229.50 |
| 2022 | Harm reduction | 1.20 | 20.58 | 72.38 | 227.44 |
| 2022 | Specialized provider | 0.31 | 19.55 | 85.76 | 251.80 |

**Supplementary Table 3: Mean total and Incremental Costs in 2022 USD and QALYs for each hepatitis C screening strategy with a time horizon to end in 2040 and 3% discount rate. The baseline, "all treatments" accounts for the number of hepatitis C treatments that occurred in Georgia to the end of 2022 within the simulation model. For each of the care pathways, the treatments due to that pathway were removed from the total treatments for each year given in the whole elimination program to model their impact, this means that the modelled QALYs are lower, and liver disease costs higher, for each strategy compared to the base case; these incremental values are negated to represent the impact of including those treatments on QALYs gained and liver disease costs averted.**

| Pathway | Cost of Diagnosis ($) compared to baseline | Cost of treatments without DAAs ($) compared to baseline | Cost of treatments with DAAs ($) compared to baseline | Liver Disease Costs ($) | Total QALYs | Incremental Costs without DAAs ($) | Incremental costs with DAAs ($) | Incremental QALYs | Base Case ICER (with DAAs)* |
|---|---|---|---|---|---|---|---|---|---|
| Baseline (all treatments) | - | - | - | 264,711,169 | 56,811,867 | - | - | - | - |
| Hospital Centralized (4,233 treatments) | 1,468,437 | 777,926 | 11,174,109 | 266,386,075 | 56,809,892 | 571,457 | 10,967,640 | 1,975 | 5752 |
| Hospital Decentralized (5,632 treatments) | 1,476,436 | 900,692 | 10,254,587 | 266,956,204 | 56,809,557 | 132,093 | 9,485,988 | 2,310 | 4258 |
| Specialized provider (4,889 treatments) | 179,229 | 906,958 | 12,594,873 | 266,713,412 | 56,809,611 | -916,056 | 10,771,859 | 2,257 | 4948 |
| HRP (469 treatments) | 24,079 | 60,148 | 726,472 | 264,924,600 | 56,811,705 | -129,204 | 537,121 | 163 | 3464 |

| | | | | | | | | | |
|---|---|---|---|---|---|---|---|---|---|
| PHC (459 treatments) | 60,891 | 75,390 | 1,016,796 | 264,915,505 | 56,811,680 | -68,055 | 873,352 | 187 | 4870 |

*ICER is mean of probabilistic ICERs; USD, United States dollars; QALYs, quality adjusted life years; DAAs, direct acting antivirals; ICER, incremental cost-effectiveness ratio; HRP, harm reduction provider; PHC, primary health care

### Supplementary Figures

**Supplementary Figure 1: (A) Cost-effectiveness plane with DAA costs; (B) cost-effectiveness plane without DAA costs for each of the screening pathways. Dotted line shows 20% of GDP per capita willingness to pay threshold, dashed line indicates 1 GDP per capita.**

**A**

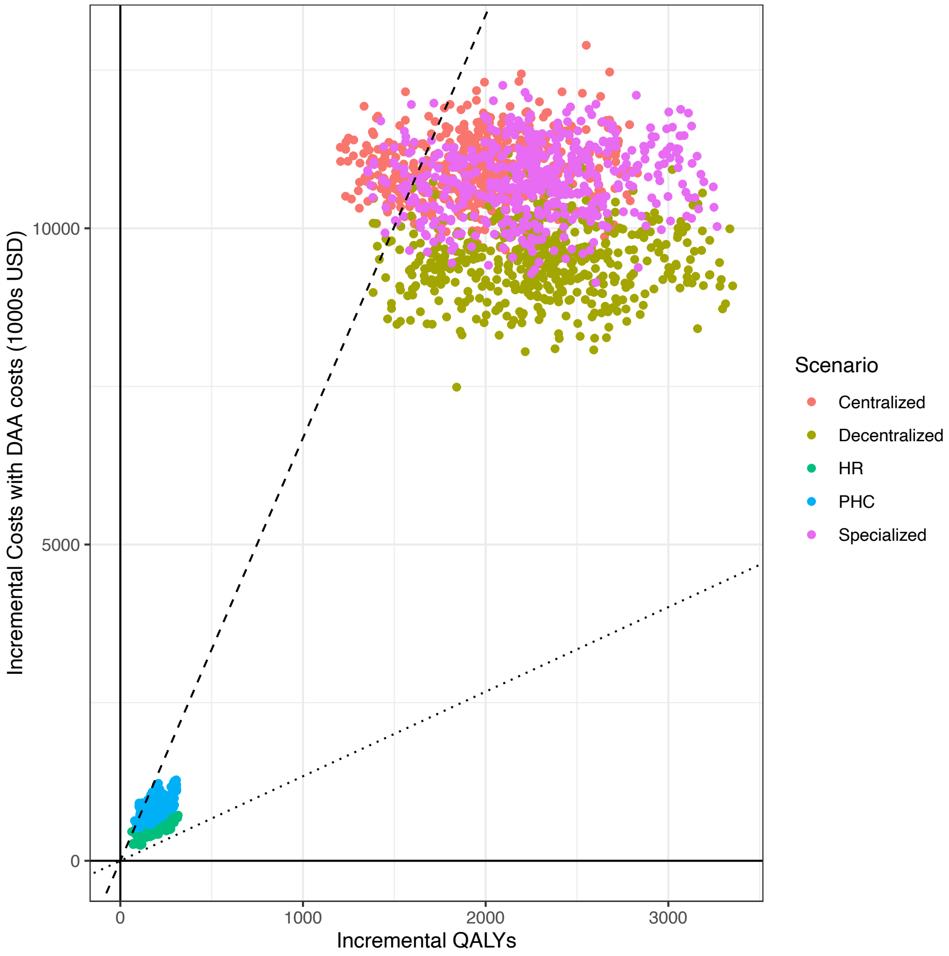

**B**

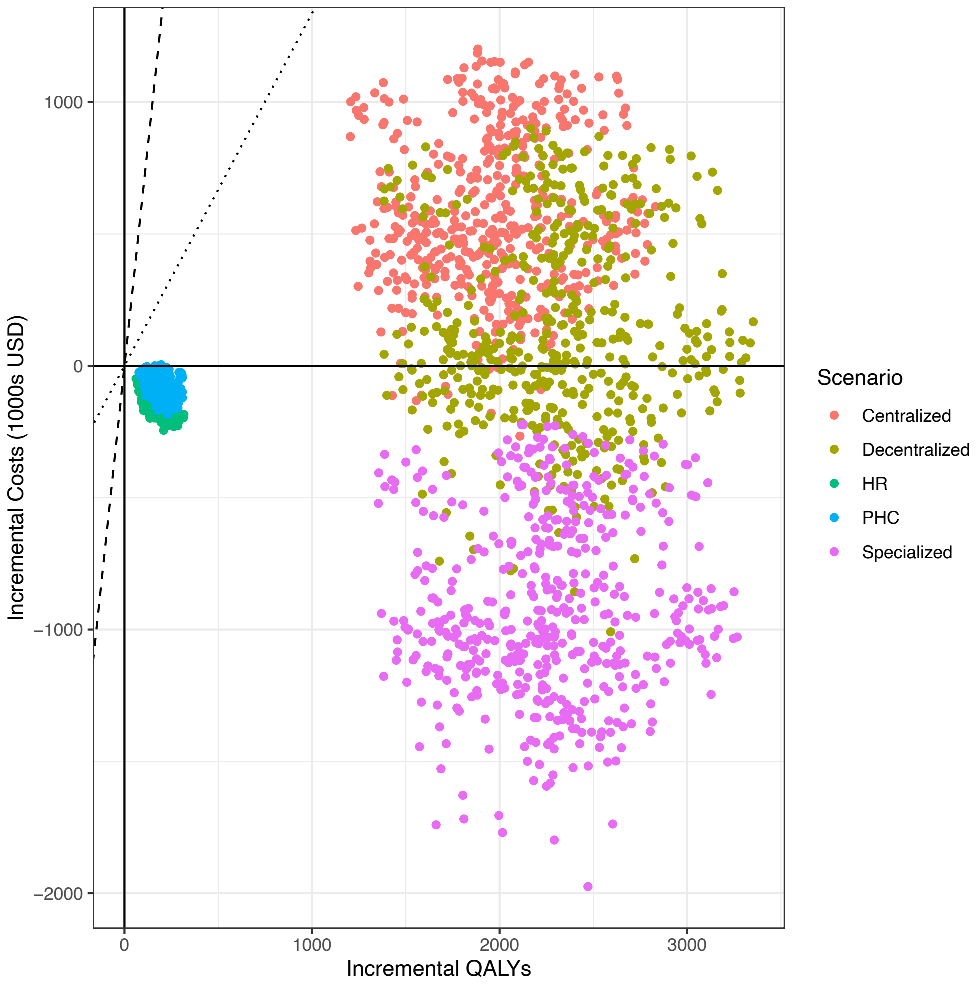


**DAA, direct acting antivirals, HR, harm reduction, PHC, primary health care.**
